# Association Between Cardiovascular Health and Pelvic Inflammatory Disease among US Adults: A Cross-Sectional Study From NHANES 2013-2023

**DOI:** 10.64898/2026.08.07.26359937

**Authors:** Gui-Yue Wang, Ying Shen, Hong Tang, Hui-Qin Mo

**Author notes:** Corresponding author: Hui-Qin Mo, Department of Obstetrics and Gynecology, The Seventh People’s Hospital of Shanghai University of Traditional Chinese Medicine, Shanghai, 200137, China. No. 358 Datong Rd. Gao Qiao, Pudong New Area Tel: 86+ 17621903598. Gui-Yue Wang and Ying Shen contributed equally to this work.

## Abstract

**Objective:** Women with a history of Pelvic Inflammatory Disease (PID) face elevated risks of health complications and mortality. This study examined the association between cardiovascular health (CVH) and PID among U.S. women.

**Methods:** We conducted a cross-sectional analysis of NHANES 2013-2023 (n=6,382). The LE8 scores were categorized into four groups based on quartiles: Q1 (<25), Q2 (25–49), Q3 (50–74) and Q4 (≥75). We calculated adjusted ORs (95% CIs) via logistic regression to evaluate LE8-PID associations.

**Results:** Compared to the highest LE8 quartile (≥75), adjusted ORs for PID were 1.66 (95%CI:1.03-2.67) for Q3, 1.71(1.10-2.67) for Q2, and 1.99(1.13-3.50) for Q1. The inverse association was consistent across health behavior and health factor components, with sleep, smoking, blood pressure, and BMI showing the strongest effects, particularly among younger women.

**Conclusions:** Higher LE8 scores are inversely associated with PID prevalence, particularly in younger women. Promoting cardiovascular health may help reduce PID burden.

## Introduction

Pelvic inflammatory disease(PID) is not only an “inflammation” but an infection-related inflammatory disorder of the upper female reproductive tract, including the endometrium, fallopian tubes, ovaries, or pelvic peritoneum(1). Among the 1,171 sexually experienced women of reproductive age surveyed in NHANES 2013-2014, the self-reported lifetime prevalence of PID was 4.4 %, indicating that an estimated 2.5 million U.S.women aged 18-44 have been diagnosed with PID during their lifetime(2). PID can lead to long-term reproductive disabilities, including infertility(3) and chronic pelvic pain(4), even closely associated with ovarian cancer(5). Individuals with PID often observed more morbidities(6), such as cardiovascular diseases (CVDs), and face a higher risk of death(7). Recurrent PID poses a significant burden on society and healthcare system. In addition to established factors such as sexual behavior, infection history, and medical procedures, recent research has include health and behavioral factors like body mass index, diet, and smoking(8, 9).

In 2010, the American Heart Association (AHA) expanded its focus from addressing existing CVDs and risk factors to strategies that could also directly promote health at the population and individual levels(10). At the core of this new approach was the creation of a new and actionable definition for the structure of cardiovascular health (CVH). The AHA introduced Life’s Simple 7 (LS7) score for assessing CVH, which takes into account seven key modifiable risk factors, including smoking, body mass index (BMI), physical activity, total cholesterol (TC), blood pressure, blood glucose, and diet. Recognizing the growing body of evidence linking sleep health with CVH, the Life’s Essential 8 (LE8) score includes sleep duration as a key component alongside the original seven metrics(11). By quantifying CVH on a scale of 0 to 100, the LE8 score provides a clearer assessment of CVH.

Previous studies have reported an association between a history of PID and an increased risk of various cardiovascular conditions, including hypertension, type 2 diabetes, myocardial infarction, and ischemic stroke(7). Conversely, those individuals with PID observed a notably reduced risk of cerebral hemorrhage(12). Given these conflicting findings, we hypothesize that PID and CVDs share common risk factors. We conducted a cross-sectional study using the NHANES database from 2013 to 2023 to investigate the association between PID and LE8.

## Materials and Methods

### Study Population

The NHANES (National Health and Nutrition Examination Survey) is a research program designed to assess the health and nutritional status of adults and children in the United States, using a population-based national cross-sectional approach. Details about the study’s design and methodologies are available at https://www.cdc.gov/nchs/nhanes/index.htm. The NHANES data set research protocol was approved by the National Center for Health Statistics Institutional Review Board, and each participant signed an informed consent form. In NHANES, surveys are conducted every two years, and each two-year period is referred to as a survey cycle. Due to the COVID-19 pandemic, the data collected from 2019 to March 2020 were integrated with the 2017-2018 cycle, thereby forming a single pre-pandemic 2017-March 2020 cycle. Moreover, the newly available data spans August 2021 to August 2023, constituting a standard two-year cycle. The NHANES data used in this study were accessed for research purposes on September 20, 2025, the date on which the final dataset was extracted. The authors analyzed only publicly available, de-identified data and did not have access to information that could identify individual participants during or after data collection. Our data were aggregated from four cycles (2013-2014, 2015-2016, 2017-March 2020, and August 2021-August 2023), which yielded a final analytical sample of 47, 639 participants. After excluding male participants(N=23, 191), women who were pregnant(N=263), those under the age of 20(N=9, 391), those missing data for LE8 score more than two metrics (N=3, 965). Ultimately, 6, 382 participants with complete information were included in the study, including 372 in the PID group and 6, 010 in the control group.

### LE8 Assessment

The LE8 scoring framework consists of eight parts, including: diet, physical activity, nicotine exposure, sleep, body mass index (BMI), non-high-density cholesterol (non-HDL), blood glucose, and blood pressure(13). The dietary metric is measured by the DASH(Dietary Approaches to Stop Hypertension) score(14), combined with total nutrient intakes obtained from the 24-hour dietary recall of study participants on the first day. Data on physical activity, nicotine exposure, sleep health, history of diabetes, and medication use are collected from self-reported questionnaires. Height, weight, and blood pressure are measured at the Mobile Examination Center (MEC). BMI is calculated as weight in kilograms divided by height in meters squared. Blood pressure is the average of three blood pressure measurements. Detailed component definitions are provided in **Supplementary Materials Table 1S.**

According to the AHA’s recommendations, the overall CVH is divided into low (LE8 score <50), medium (LE8 score ≥50 but <80), and high (LE8 score ≥80) levels(13). Consequently, the study participants were divided into four groups: Quartile 1 (LE8 <25), Quartile 2 (25 ≤ LE8 <50), Quartile 3 (50 ≤ LE8 <75), and Quartile 4 (LE8 ≥75).To investigate and emphasize the impacts of lifestyle interventions on PID, we categorized LE8 into two distinct groups: health factors (BMI, non-HDL cholesterol, blood glucose, and BP) and behavior factors (diet, physical activity, nicotine exposure, and sleep).

### Pelvic inflammatory disease Assessment

The history of PID is assessed using RHQ-078. The questionnaire asks: “Have you/Has SP ever been treated for an infection in your/her fallopian tubes, uterus, or ovaries, also called a pelvic infection, pelvic inflammatory disease, or PID?” Value descriptions include: yes, no, refused, don’t know, and missing.

### Covariate Assessment

Important covariates were included: age, race, education level, family income, and sexual behavior characteristics. The specific categorizations were as follows: age; race/ethnicity (Mexican American, Non-Hispanic Black, Non-Hispanic White, Other Hispanic, Other Races); education level (less than high school, high school, some college or above); poverty-income ratio (<1.5, 1.5-4.99, ≥5.0); and age at first sexual intercourse, always condom use (yes/no), and history of sexually transmitted infections (STIs, yes/no).

### Statistical Analysis

The data were examined for missing and for logic checks in accordance with CDC recommendations. Missing data on covariates were imputed with the median value based on the surrounding three data points using IBM SPSS Statistic. Consistent with CDC guidelines (https://wwwn.cdc.gov/nchs/nhanes/tutorials/weighting.aspx), the complex, multi-stage cluster survey design of NHANES was accounted for in all statistical analyses by applying the appropriate sampling weights. We constructed a unified analysis weight for the pooled dataset. The base Full Sample 2 Year MEC Exam Weight (WTMEC2YR) was first scaled by the duration of its respective cycle.

The data weight of 2017-March 2020 was obtained by dividing the Full sample MEC exam weight (WTMECPRP) by 3.2, while for other cycles, the WTMEC2YR was divided by 2. Subsequently, all scaled weights were normalized to sum to the total sample size. Descriptive statistics were presented using mean values with standard deviations (SDs) for continuous data, and using numbers and percentages for categorical variables. Categorical variables were analyzed using a weighted chi-square test, while the weighted two-samples Student’s t-test was employed for variables that follow normal distribution and one-way analysis of variance (ANOVA) for multiple groups. The study employed weighted logistic regression models to calculate odds ratios (ORs) and 95% confidence intervals (CIs) in order to assess the association between cardiovascular health, represented as continuous LE8 scores or categorical quartiles of LE8 scores, and the history of pelvic infection (yes vs no). We have evaluated multicollinearity using variance inflation factors (all VIFs < 2.0 indicating no substantial multicollinearity). Besides, adjusted ORs were obtained after adjusting confounding factors: age, race, STIs, age at first sexual intercourse and education level and family income. Additional analyses using survey-weighted modified Poisson regression with robust variance to estimate prevalence ratios (PRs) was conducted. To model the association between continuous CVH and ORs of PID, 4-knots (5th, 35th, 65th, 95th) restricted cubic splines (RCS) was performed using software package R, with confounding covariates adjusted. The subgroup analyses were exploratory and post-hoc, aimed at assessing whether the association differs across categories of covariates. Analyses were stratified by the following covariates: age (<50 vs. ≥50 years), education level (less than high school, high school, or some college or above), family income-to-poverty ratio (<1.5, 1.5-4.99, ≥5.0), history of STIs (yes/no), age at first sexual intercourse ≥17 years (yes/no), and race/ethnicity (Mexican American, non-Hispanic Black, non-Hispanic White, Other Hispanic, and Other Races).

Interaction effects were assessed using likelihood ratio tests, comparing the model with the interaction term against the nested model without the term. The weighted logistic regression was also conducted to evaluate the relationship between the single metric with PID. For this analysis, OR for the overall LE8 score was calculated for each 10-point increase. Two distinct models were constructed for each component: 1) to estimate the OR for PID per 10-point increase in the component score, and 2) to estimate the OR by comparing participants with component scores above the average to those with scores below the average. All analyses were adjusted for age, education level, family income, STIs, age at first sexual intercourse and race. A Bonferroni correction was applied to adjust for multiple comparisons across groups. All analyses were performed using RStudio software (version 4.3.3; Posit Software, PBC). P-values of < 0.05 were considered statistically significant, using a two-sided test.

## Results

### Baseline characteristics of our study participants

A total of 6,382 women met the inclusion criteria and were analyzed. **Fig 1** shows the flowchart of the study population. **Table 1** shows the differences in weighted baseline characteristics of all participants, including age, ethnicity, education, ratio of family income to poverty, age at first sexual intercourse, condom use, STIs history and relevant LE8 metrics. After accounting for the complex survey design weights (representing a national population of 117,754,136), the analytical weighted sample comprised 323 participants with PID and 6, 069 controls. Based on the weighted analysis, we observed significant differences between PID patients and the control group. Participants with older age, Non-Hispanic White, and lower education level and family income had higher rates of PID. Individuals with PID were more likely to have an earlier sexual debut and a reported history of STIs. The LE8 score was lower in PID patients than controls (63.6 ±14.6 vs. 70.0 ± 15.2). For the CVH metrics, there were differences between the two groups in nicotine exposure, sleep health, BMI, blood lipids and blood pressure (p < 0.05). However, there is no significant difference observed in DASH, physical activity and blood glucose between two groups.

**Fig 1.**
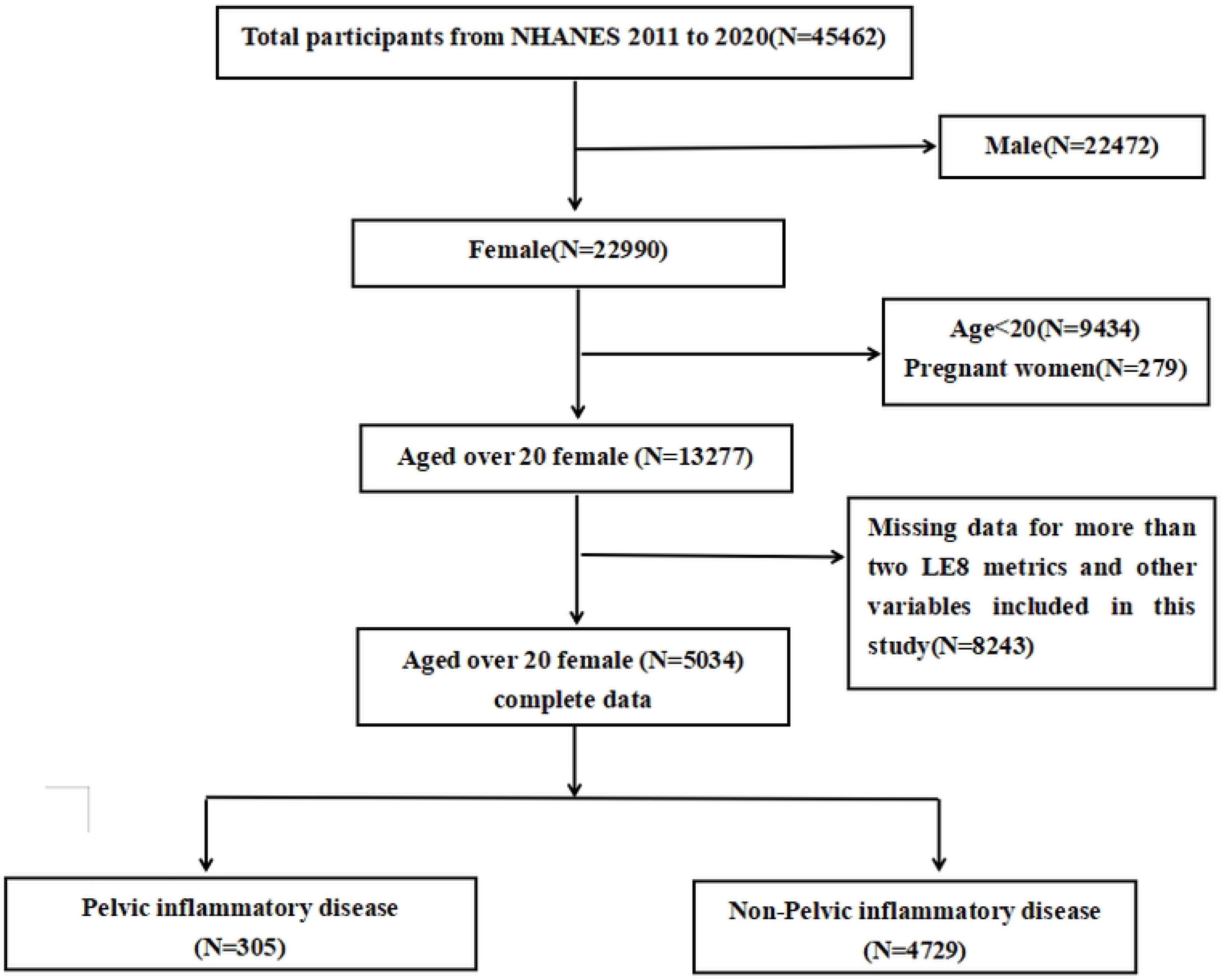
Flowchart of the study population.

**Table 1.**
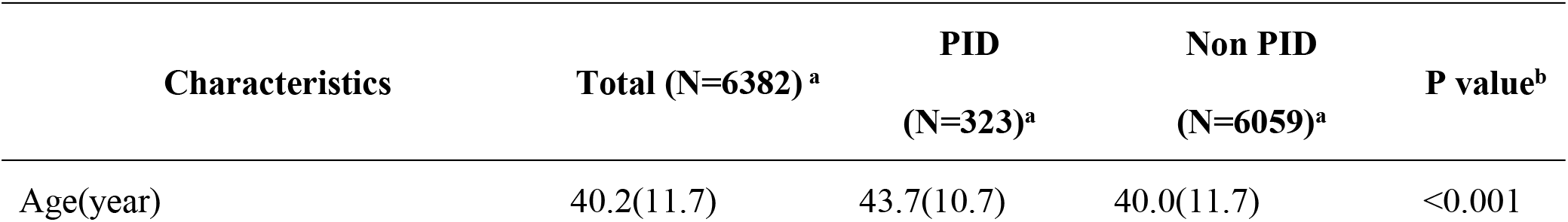
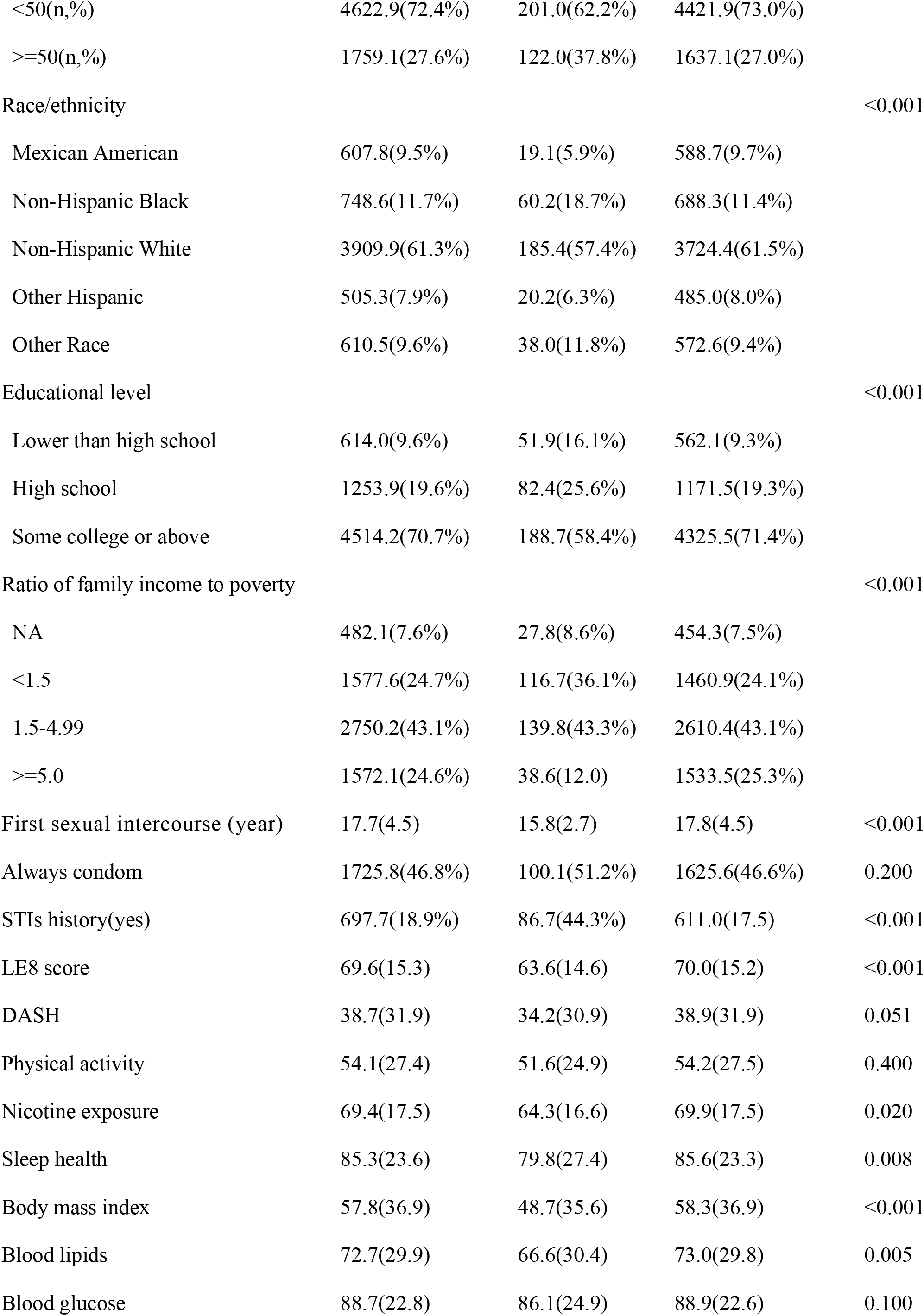
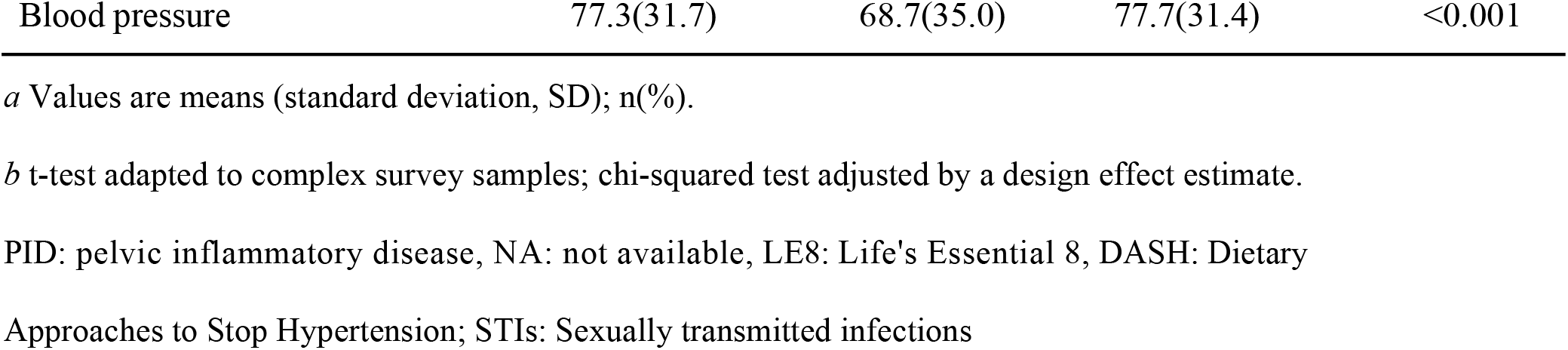
NHANES 2013-2023 Baseline Characteristics by Pelvic Inflammatory Disease (Complex Survey Design Weighted Analysis, Weighted sample= 117,754,136)

| Characteristics | Total (N=6382)<sup>a</sup> | PID<br>(N=323)<sup>a</sup> | Non PID<br>(N=6059)<sup>a</sup> | P value<sup>b</sup> |
| --- | --- | --- | --- | --- |
| Age(year) | 40.2(11.7) | 43.7(10.7) | 40.0(11.7) | <0.001 |
| <50(n,%) | 4622.9(72.4%) | 201.0(62.2%) | 4421.9(73.0%) |  |
| >=50(n,%) | 1759.1(27.6%) | 122.0(37.8%) | 1637.1(27.0%) |  |
| Race/ethnicity |  |  |  | <0.001 |
| Mexican American | 607.8(9.5%) | 19.1(5.9%) | 588.7(9.7%) |  |
| Non-Hispanic Black | 748.6(11.7%) | 60.2(18.7%) | 688.3(11.4%) |  |
| Non-Hispanic White | 3909.9(61.3%) | 185.4(57.4%) | 3724.4(61.5%) |  |
| Other Hispanic | 505.3(7.9%) | 20.2(6.3%) | 485.0(8.0%) |  |
| Other Race | 610.5(9.6%) | 38.0(11.8%) | 572.6(9.4%) |  |
| Educational level |  |  |  | <0.001 |
| Lower than high school | 614.0(9.6%) | 51.9(16.1%) | 562.1(9.3%) |  |
| High school | 1253.9(19.6%) | 82.4(25.6%) | 1171.5(19.3%) |  |
| Some college or above | 4514.2(70.7%) | 188.7(58.4%) | 4325.5(71.4%) |  |
| Ratio of family income to poverty |  |  |  | <0.001 |
| NA | 482.1(7.6%) | 27.8(8.6%) | 454.3(7.5%) |  |
| <1.5 | 1577.6(24.7%) | 116.7(36.1%) | 1460.9(24.1%) |  |
| 1.5-4.99 | 2750.2(43.1%) | 139.8(43.3%) | 2610.4(43.1%) |  |
| >=5.0 | 1572.1(24.6%) | 38.6(12.0) | 1533.5(25.3%) |  |
| First sexual intercourse (year) | 17.7(4.5) | 15.8(2.7) | 17.8(4.5) | <0.001 |
| Always condom | 1725.8(46.8%) | 100.1(51.2%) | 1625.6(46.6%) | 0.200 |
| STIs history(yes) | 697.7(18.9%) | 86.7(44.3%) | 611.0(17.5) | <0.001 |
| LE8 score | 69.6(15.3) | 63.6(14.6) | 70.0(15.2) | <0.001 |
| DASH | 38.7(31.9) | 34.2(30.9) | 38.9(31.9) | 0.051 |
| Physical activity | 54.1(27.4) | 51.6(24.9) | 54.2(27.5) | 0.400 |
| Nicotine exposure | 69.4(17.5) | 64.3(16.6) | 69.9(17.5) | 0.020 |
| Sleep health | 85.3(23.6) | 79.8(27.4) | 85.6(23.3) | 0.008 |
| Body mass index | 57.8(36.9) | 48.7(35.6) | 58.3(36.9) | <0.001 |
| Blood lipids | 72.7(29.9) | 66.6(30.4) | 73.0(29.8) | 0.005 |
| Blood glucose | 88.7(22.8) | 86.1(24.9) | 88.9(22.6) | 0.100 |
| Blood pressure | 77.3(31.7) | 68.7(35.0) | 77.7(31.4) | <0.001 |

*a* Values are means (standard deviation, SD); n(%).
*b* t-test adapted to complex survey samples; chi-squared test adjusted by a design effect estimate.
PID: pelvic inflammatory disease, NA: not available, LE8: Life's Essential 8, DASH: Dietary
Approaches to Stop Hypertension; STIs: Sexually transmitted infections

### Associations of LE8 score with PID Risk

**Table 2** shows the association of different LE8 score categories with PID risk. In multivariate logistic regression, compared with Q4, the adjusted ORs for PID in Q3, Q2, and Q1 were 1.66 (95% CI: 1.03-2.67), 1.71 (95% CI: 1.10-2.67), and 1.99 (95% CI: 1.13-3.50), respectively. Prevalence ratio analyses yielded comparable results. A significantly increased PID risk was also observed across lower quartiles of health behaviors and health factors, with Q1 showing the highest risk, consistent with the overall CVH findings.

**Table 2.** Survey-Weighted Associations of Life’s Essential 8 score with PID Risk.

| Variables | PID(n,%) | Unadjusted<sup>a</sup><br>OR(95%CI) | Adjusted<sup>b</sup><br>OR(95%CI) | Adjusted<sup>b</sup><br>PR(95%CI) |
| --- | --- | --- | --- | --- |
| Quartiles of LE8 score |  |  |  |  |
| Quartile 1 | 117(7.8) | 3.27 (1.97-5.45) | 1.99 (1.13-3.50) | 1.92 (1.12-3.28) |
| Quartile 2 | 84(5.5) | 2.25 (1.49-3.41) | 1.71 (1.10-2.67) | 1.68 (1.10-2.56) |
| Quartile 3 | 78(4.8) | 1.96 (1.23-3.13) | 1.66 (1.03-2.67) | 1.63 (1.03-2.56) |
| Quartile 4 | 44(2.5) | 1(ref) | 1(ref) | 1(ref) |
| P for trend <sup>c</sup> |  | <0.001 | <0.001 |  |
| Quartiles of health behaviors score |  |  |  |  |
| Quartile 1 | 93(6.7) | 1.82 (1.18-2.80) | 1.45 (0.91-2.32) | 1.42 (0.91-2.20) |
| Quartile 2 | 88(5.4) | 1.45 (0.88-2.39) | 1.40 (0.85-2.30) | 1.37 (0.86-2.19) |
| Quartile 3 | 77(4.6) | 1.22 (0.79-1.90) | 1.16 (0.74-1.80) | 1.15 (0.76-1.74) |
| Quartile 4 | 65(3.8) | 1(ref) | 1(ref) | 1(ref) |
| P for trend <sup>c</sup> |  | <0.001 | 0.015 |  |

| Quartiles of health factors score |  |  |  |  |
| --- | --- | --- | --- | --- |
| Quartile 1 | 106(7.1) | 2.85 (1.69-4.81) | 1.76 (0.98-3.15) | 1.71 (0.98-2.98) |
| Quartile 2 | 89(6.2) | 2.43 (1.54-3.84) | 1.80 (1.11-2.92) | 1.76 (1.11-2.78) |
| Quartile 3 | 83(4.7) | 1.82 (1.05-3.14) | 1.53 (0.86-2.71) | 1.51 (0.87-2.61) |
| Quartile 4 | 44(2.6) | 1(ref) | 1(ref) | 1(ref) |
| P for trend <sup>c</sup> |  | <0.001 | 0.003 |  |

*a* unadjusted
*c* P for trends were performed by modeling the quartiles as a continuous variable (scores 1-4).
PID: pelvic inflammatory disease; CI: confidence interval; OR: odds ratio; PR: prevalence ratio

To assess the continuous association between PID risk and the LE8 scores, we used the restricted cubic splines (RCS) in **Fig 2**. The graph visually shows the linear relationship indicating a trend of decreasing PID risk as LE8 increases, as well as in health behaviors(**Fig 2B**) and health factors group(**Fig 2C**). In the health factors(**Fig 2C**), there was a linear decreasing trend in the prevalence of PID as the score increased, with no evidence of nonlinearity (P=0.585). All RCS models were adjusted for the full covariate set.

**Figure 2.**
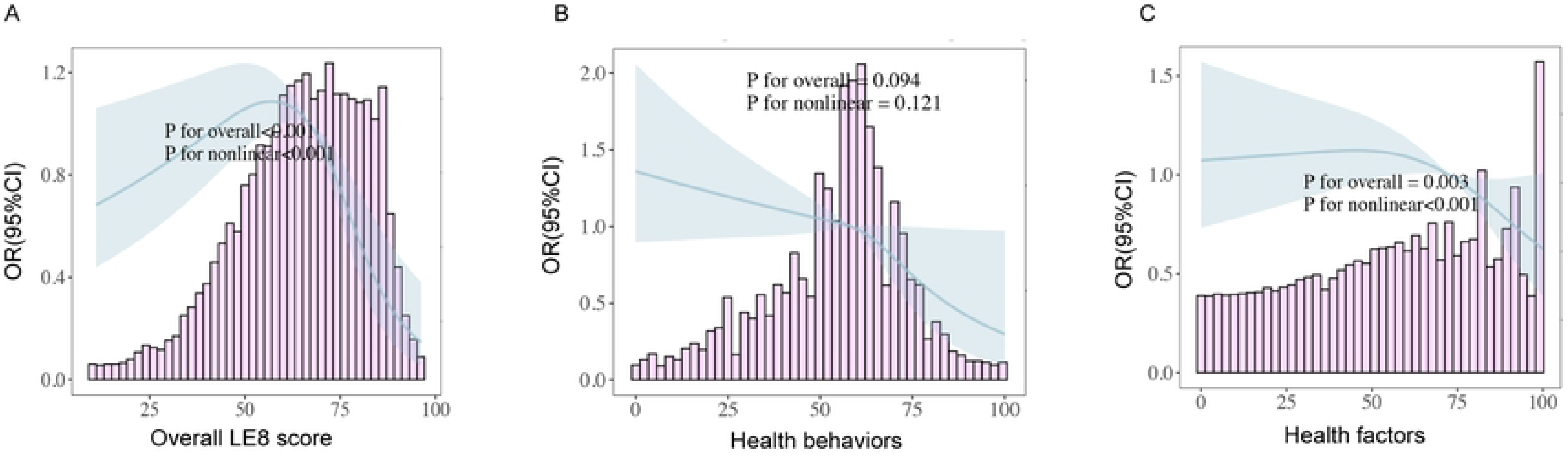
Dose–response relationships of PID with Life’s Essential 8 scores, health behavior scores, and health factors scores. Legend: The RCS represent adjusted ORs and 95% CIs of overall CVH scores in relation to PID(Figure 2a), as well as the score of health behaviors (Figure 2b) and the health factors scores (Figure 2c). The histogram represents frequency of participants according to CVH scores. The solid line represents point estimates of ORs, and the shaded areas represent the 95%CI. ORs were calculated using logistic regression and were adjusted for age (continuous), education level (less than high school, high school, some college or above), family income (<1.5, 1.5-4.99, ≥5.0), sexually transmitted infection (yes/no), age at first sexual intercourse (continuous) and race (Mexican Americans, non-Hispanic blacks, non-Hispanic whites, other Hispanics, other races).

### Subgroup analysis of each LE8 metric in relation to PID risk

The forest plot **Fig 3** provides subgroup analysis, indicating that the association between LE8 metrics and PID varies in different age categories, ethnicity, education levels, family income, age at first sexual intercourse and STIs history. A generally consistent association between LE8 and reduced PID prevalence was observed across subgroups, with significant interaction effects only for age and race. The strength of this association varied markedly, being most robust among participants under 50 years old (OR = 0.764, 95% CI: 0.693-0.844; P = 0.01, **Fig 3**) and the “Other Race” group (OR = 0.545, 95% CI: 0.424-0.691). This suggests that the protective effect of optimal cardiovascular health is particularly significant in these populations.

**Figure 3.**
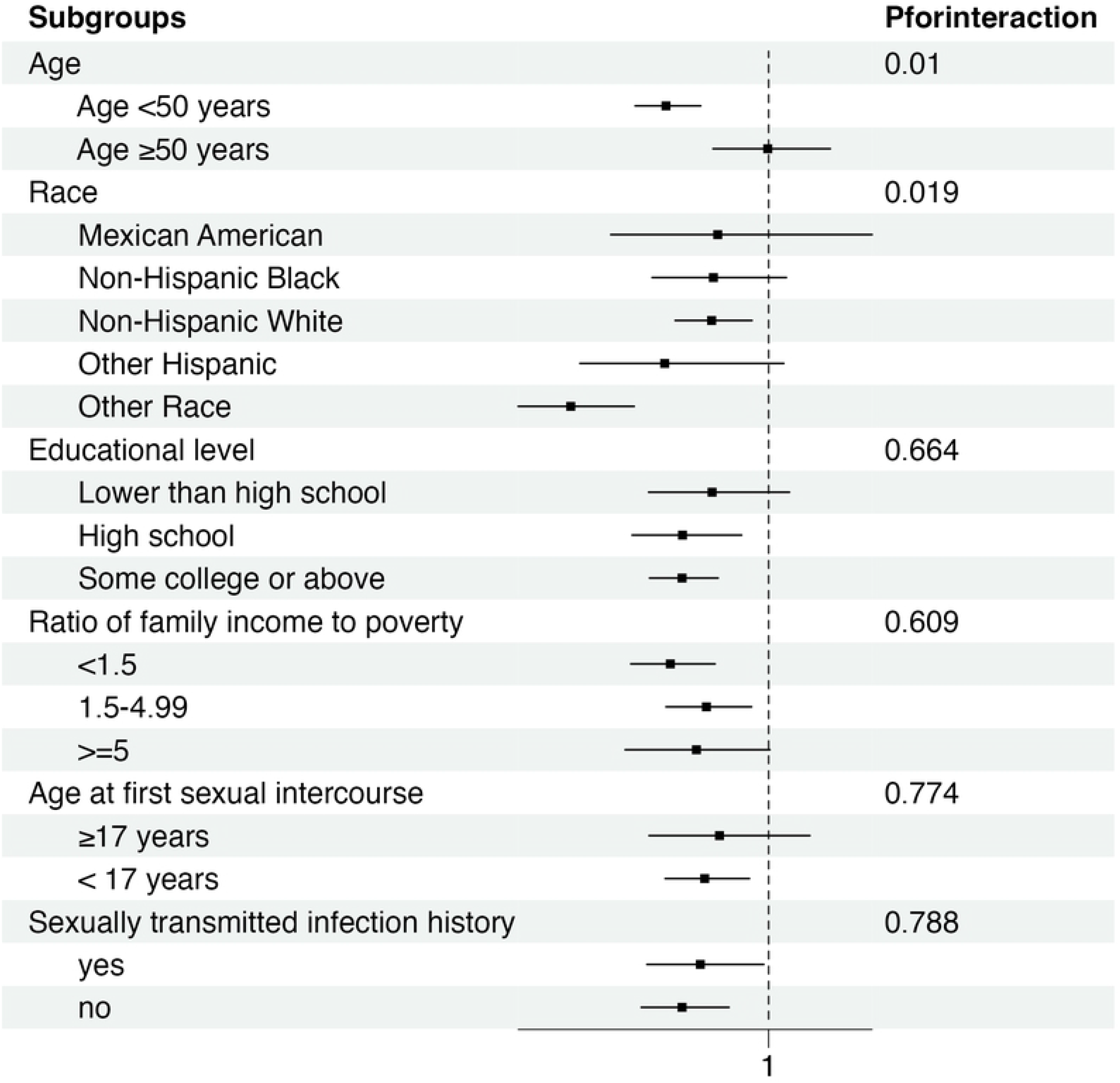
Subgroup analysis of the association of the Life’s Essential 8 scores and PID. Legend: ORs were calculated as each 10 points increase in LE8 score. Each stratification was adjusted for age (continuous), education level (less than high school, high school, some college or above), family income (<1.5, 1.5-4.99, ≥ 5.0), sexually transmitted infection (yes/no), age at first sexual intercourse (continuous) and race (Mexican Americans, non-Hispanic blacks, non-Hispanic whites, other Hispanics, other races).

As shown in **Fig 4**, each health behavior metric was independently associated with lower PID risk, with diet showing the smallest effect per 10-point increase (OR = 0.84; 95%CI: 0.56–1.17), while no significant associations were observed for health factors (blood lipids, blood pressure, BMI). Compared with lower scores, the nicotine exposure ≥ 60 group had the largest risk reduction (OR = 0.38; 95%CI: 0.20–0.73), followed by sleep health ≥ 70 group (OR = 0.61; 95%CI: 0.62–0.90).

**Figure 4.**
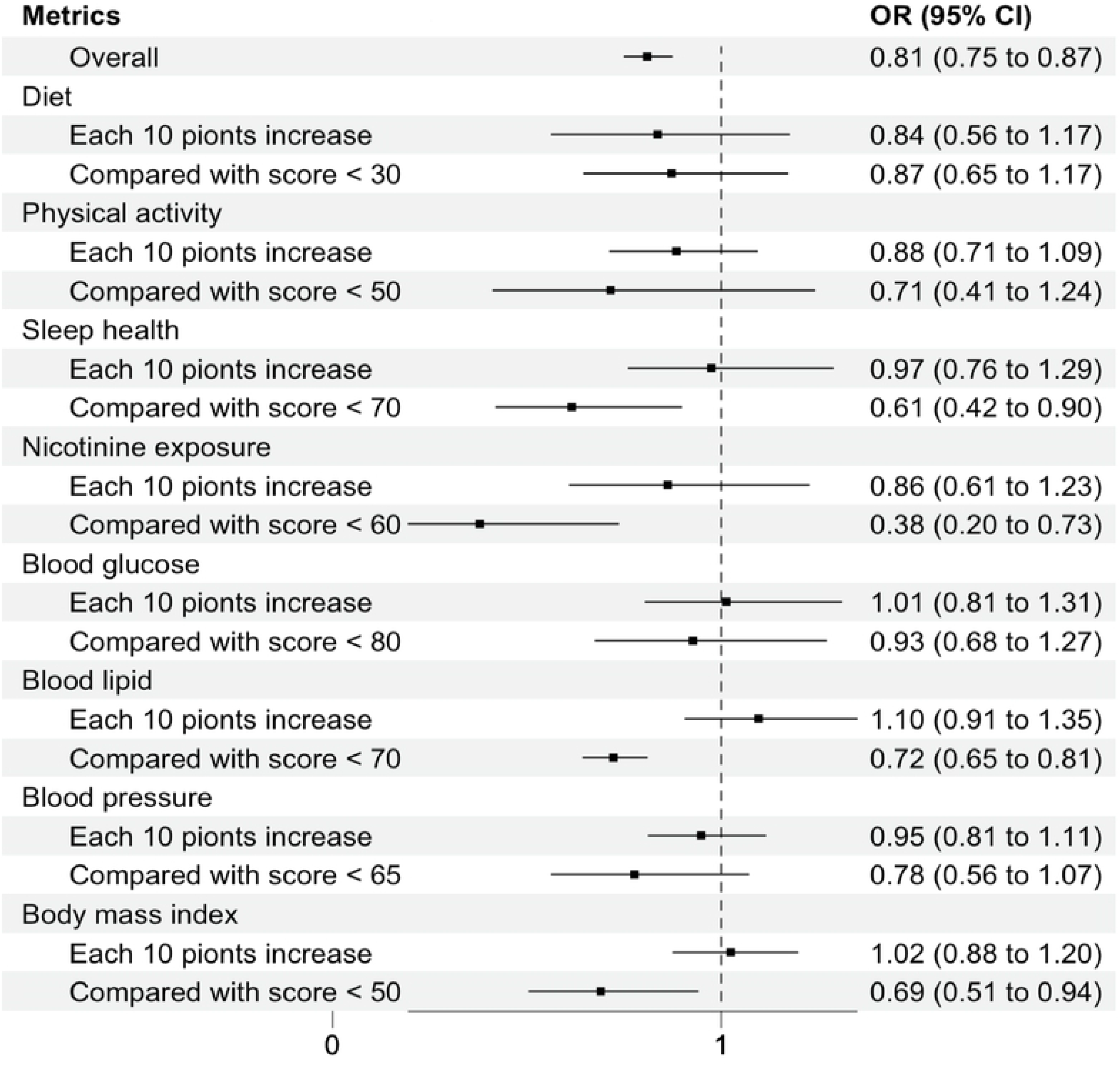
Adjusted ORs of each Life’s Essential 8 metrics with PID risk. Legend: Each metric was adjusted for age (continuous), education level (less than high school, high school, some college or above), family income (<1.5, 1.5-4.99, ≥5.0), sexually transmitted infection (yes/no), age at first sexual intercourse (continuous) and race (Mexican Americans, non-Hispanic blacks, non-Hispanic whites, other Hispanics, other races).

## Discussion

Our study found that women with a higher LE8 is significantly correlated with a decrease risk of PID. The results of its sub-scales of health behaviors and health factors remain consistent and robust in subgroup and sensitivity analysis. Among the 8 LE8 metrics, nicotine exposure, sleep health, BMI, blood lipids and blood pressure are identified as the main contributors to the negative correlation between LE8 scores and PID.

For the individual metrics, we found that nicotine exposure was the major individual contributor to reduced PID prevalence. A regression study showed that, compared with never-smoking women, current smokers had a significantly elevated relative risk of PID(15). A North Queensland cross-sectional study revealed that young women with PID had prolonged hospital stays, strongly associated with smoking(8). Nicotine and related compounds can impair mucosal immunity and ciliary function in the fallopian tubes, facilitating infection(16). Sleep health was newly included in the LE8 in 2022(11). But current few research indicates that sleep health may be a risk factor of PID. PID may increase the risk of depressive disorder, anxiety disorder and sleep disorder, which will impair life quality(17). Mechanistically, sleep disturbance causing neuroinflammation and oxidative stress(18) influence the occurrence and development of PID requires further investigation. It is worth noting that there were no statistically significant differences in physical activity and DASH between the PID and non-PID groups. Dietary mineral intake such as copper and magnesium, which have been associated with PID(9) .

Individuals with significantly decreased health factor scores are typically considered to have one or more of the following conditions: diabetes, hypertension, hyperlipidemia, and higher body weight. Okoth et al showed that PID is associated with an increase in hypertension and type 2 diabetes, two major risk factors for CVDs(7). In addition, Liou TH et al. showed that chronic inflammatory response caused by PID can lead to atherosclerosis, thereby increasing the risk of CVDs(19). Besides, long-term use of antibiotics for PID may cause QT interval prolongation and torsades de pointes(20), increasing malignant arrhythmias risk.

For the LE8 sub-scales, we found that health factors and behaviors were inversely associated with PID. We speculate that modifiable behaviors, which include smoking, diet, physical activity, and sleep, may directly impact PID risk in the shorter term by modulating inflammation and immune function. Promoting these healthy behaviors not only improves intermediary health factors like obesity, blood lipids, glucose, and blood pressure but may also concurrently mitigate the risk of developing PID.

This observed relationship may be explained by several underlying biological mechanisms. Smoking, physical inactivity, poor diet, and insufficient sleep not only impair CVH but also weaken genital-tract immune defenses and increase exposure to STIs and the PID(21). Most cases of PID are caused by *Chlamydia trachomatis*, this pathogen can be detected in cardiovascular tissues such as arterial walls and heart valves. Through local infiltration and persistent low-grade inflammation, it directly damages the vascular endothelium and initiates the atherosclerotic process, thereby creating a direct physical link between PID and impaired CVH(7). The acute phase of PID releases large amounts of pro-inflammatory cytokines such as IL-6, TNF-α, and IL-1β, which induce insulin resistance, increase oxidative stress, inhibit eNOS, lead to endothelial dysfunction, promote LDL oxidation and macrophage uptake, and accelerate atherosclerotic plaque formation(22). Further research using prospective cohorts with acute PID diagnoses and serial biomarker measurements is needed to clarify the underlying mechanisms.

Our study indicates that LE8 and lower PID prevalence was consistent particularly noticeable in the participants under 50 years old and a healthy lifestyle is more meaningful for improving CVH in young women with PID. During the menopausal transition, declining oestrogen and progesterone reduce insulin sensitivity, redistribute body fat, alter lipids (higher LDL, lower HDL), impair endothelial function and amplify inflammation, thereby increasing not only cardiometabolic risk(23) but also pelvic inflammatory disease(24). Studies have revealed that the highest self-reported frequency of PID treatment among women earning below 150% of the federal poverty level and without a high school education(25). Income, wealth, and socioeconomic instability are well-established social determinants of health. They significantly influence physical and mental well-being by increasing the risk of conditions such as sleep disturbances, hypertension, and diabetes. Low educational attainment predicts poorer employment prospects and economic insecurity, which in turn obstruct healthy lifestyles and access to health care—All of the above are risk factors for PID. Effective public health interventions should target structural health inequities while promoting cardiovascular health to reduce the burden of PID.

At present, few articles have examined the relationship between PID and LE8. Tong et al. first used NHANES data to illustrate this association(21). In contrast, our study extends their findings by using a larger and more recent sample (NHANES 2013-2023), emphasizing the dose-response relationship. Additionally, we separately examined health behavior and health factor subscales, conducted subgroup analyses on individual LE8 metrics, and comprehensively adjusted for demographic and behavioral confounders. These enhancements improve the robustness and interpretability of our findings.

This article has certain limitations. Due to the nature of this article as a cross-sectional study, it cannot establish causality. Self-reported questionnaires for assessing health behavior may introduce recall bias, as participants may not accurately recall or report their behavior. At the same time, the severity of PID was not considered. Although we adjusted for a wide range of potential confounders in our multivariate analysis, the possibility of residual confounding due to unmeasured or unknown factors like medical procedures cannot be entirely ruled out. Some LE8 components, such as physical activity and sleep, had relatively high missing data which may lead to potential bias. Prospective cohort studies and randomized trials targeting LE8-related interventions are warranted to clarify the relationship between CVH and PID.

## Conclusions

In summary, this nationally representative study of U.S. adults found that cardiovascular health assessed by LE8 scores is negatively correlated with PID. Achieving ideal cardiovascular health through lifestyle modifications may help reduce PID risk, thereby lessening the burden on public health efforts.

## Data Availability

Data from the National Health and Nutrition Examination Survey (NHANES) 2011–2023 are publicly available online (https://www.cdc.gov/nchs/nhanes/index.htm).

https://github.com/xiangwenkai/cardiovascular_pelvic

## Acknowledgements

This research did not receive any specific grant from funding agencies in the public, commercial, or not-for-profit sectors. The authors acknowledge gratitude to all the staff who participated in this study. We would thank Wen-Kai Xiang for his guidance on the statistics.

## Authors’ contributions

H.M. and G.W. conceived and designed the study. H.M. and G.W. performed data acquisition and analysis. H.M., G.W., Y.S., and H.T. contributed to manuscript preparation. H.M. assumed primary responsibility for final content integrity. All authors critically reviewed and approved the final manuscript.

## Availability of data and materials

Data from the National Health and Nutrition Examination Survey (NHANES) 2011–2023 are publicly available online (https://www.cdc.gov/nchs/nhanes/index.htm). We provide a public GitHub repository for the LE8 construction (https://github.com/xiangwenkai/cardiovascular_pelvic).

## Ethics approval and consent to participate

This study utilized publicly available, de-identified data from the NHANES cycles spanning 2013-2014, 2015-2016, 2017-March 2020, and August 2021-August 2023. The NHANES study protocols were reviewed and approved by the National Center for Health Statistics (NCHS) Ethics Review Board. The specific protocol numbers for the cycles included in our analysis are as follows: Continuation of Protocol #2011-17 (for 2013-2014, 2015-2016, and the portion of 2017-March 2020 prior to October 26, 2017), Protocol #2018-01 (for the portion of 2017-March 2020 on and after October 26, 2017), and Protocol #2021-05 (for August 2021-August 2023) . All original participants provided written informed consent. As this study involves only secondary analysis of pre-existing, publicly available, and fully anonymized data without any direct contact with human subjects, it was determined to be exempt from additional institutional review board approval.

## List of abbreviations

AHA: American Heart Association
ANOVA: analysis of variance
BMI: body mass index
CVD: cardiovascular diseases
CVH: cardiovascular health
CIs: confidence intervals
DASH: Dietary Approaches to Stop Hypertension
LS7: Life’s Simple 7
LE8: Life’s Essential 8
MEC: Mobile Examination Center
non-HDL: non-high-density lipoprotein
NHANES: National Health and Nutrition Examination Survey
SEs: standard errors
RCS: restricted cubic splines
PID: Pelvic Inflammatory Disease

